# Early indicators of child obesity to aid future clinical trials for lifecycle obesity prevention

**DOI:** 10.64898/2026.05.13.26353150

**Authors:** Carol A. Wang, Kristin L. Connor, Sadra Mohammadkhani, Stephen J. Lye, Trevor A. Mori, Lawrence J. Beilin, Craig E. Pennell

**Affiliations:** School of Medicine and Public Health, University of Newcastle, New South Wales, Australia; Mothers’ and Babies’ Research Program, Hunter Medical Research Institute, New South Wales, Australia; Department of Health Sciences, Carleton University, Ottawa, Canada; Alliance for Human Development, Lunenfeld-Tanenbaum Research Institute, Toronto, Canada; Medical School, Royal Perth Hospital Unit, University of Western Australia, Western Australia, Australia

**Keywords:** Early predictors, Genetics, Obesity, Preventive Medicine

## Abstract

**Background:** 39M children worldwide are overweight or have obesity, accelerating risk for adult non-communicable diseases. Presently, interventions to prevent obesity have had limited success due to poor timing and lack of personalisation.

**Objective:** We aimed to identify early-life predictors of childhood obesity (ChOB) that could aid targeting specific population subsets for obesity prevention interventional studies.

**Methods:** Data were from the Raine Study Gen2 participants (n=1494). Anthropometric and genetic predictors evaluated included birthweight (BW), early-life BMI (1-3 years), and three polygenic scores (PGS) [two BW-PGSs (BW-PGS_2016_ and BW-PGS_2019_) and a ChOB-PGS], developed from BW and ChOB genome-wide-association-studies, respectively. Multivariate analyses were performed to investigate associations between predictors and child-BMI (5-, 8- , 10-years).

**Results:** BW-PGS_2019_ associate with child-BMI at 5-years. BW-PGS_2016_ was not associated with child-BMI. Remaining predictors positively associate with child-BMI at 5-, 8- and 10-years (p<0.001). Early-life BMI, ChOB-PGS and BW accounted for up to 38.7%, 5.8% and 3.4% of the variability in child-BMI, respectively.

**Conclusions:** Our data suggest early-life BMI is a better predictor of child-BMI than ChOB-PGS, and BW, accounting for up to ten-fold more variance in child-BMI. Future interventional studies to mitigate obesity could target early-life BMI as a marker to identify children at the highest risk.

**Practitioner Points (up to 3 key points):** 1. Birthweight is associated with later-life obesity, but it is a variable predictive marker. Polygenic scores (PGS) provide additional insight into obesity and overweight risk in adulthood. Little is known about other predictors that could provide insight into the risk of later-life obesity.

2. Early-life BMI (before 3-years of age) predicts childhood obesity better than genetic and birthweight markers, accounting for more variance. Interventions to prevent obesity could now be more appropriately targeted.

3. Early-life BMI is a better predictor of child BMI than birthweight or childhood obesity PGS, which could aid obesity risk-stratification and ensure interventions to prevent obesity are targeted to children who will most benefit.

## Introduction

Thirty-nine million children worldwide are overweight or have obesity^1^, accelerating risk for adult non-communicable diseases^2,3^. Although birthweight (BW) has been shown to predict risk for later obesity^4^, a focus on BW alone has delayed progress in identifying which subset of children are at greatest risk for obesity earlier in the lifecycle^5,6^. Additionally, few studies have considered multiple anthropometric markers of child obesity to determine which have the greatest predictive power. Collectively, these gaps are critical to close if we aim to prevent obesity at the earliest possible stages.

Birthweight polygenic scores (BW-PGS) have provided additional insight into the roles of genes in adult overweight and obesity risk, revealing subgroups of individuals at low and high relative risk for obesity^7^. Previous studies using Raine Study data demonstrated associations between obesity risk genetic variants and BMI trajectories in childhood^8,9^. Thus, genetic tools, including PGS, could accelerate opportunities to prevent and manage obesity in both childhood and adulthood^8–10^. However, it remains unknown if these genetic tools better predict pathways to obesity than other clinically-accessible markers^10,11^. Additionally, it is unknown if anthropometric and genetic markers stably predict BMI trajectory across child and adolescent years. This information is critical to uncover which individuals are at greatest risk for early high BMI and at which time, a key first step to target early interventions with the greatest potential to prevent child obesity.

We aimed to identify early life predictors of childhood obesity that could aid in identifying specific population subsets for future obesity prevention interventional studies. This study used data from the Raine Study, Perth, Western Australia, Gen2 study participants with genetics and anthropometric data available for analyses. We hypothesised that i. early BMI would better predict child obesity at 5-, 8- and 10-years of age than birthweight, ii. early genetics would also predict child obesity, and iii. the robustness of the predictions would depend on age at assessment.

## Methods

### Study Population

The Raine Study is a prospective pregnancy cohort that recruited 2900 women at Western Australia’s major perinatal centre, King Edward Memorial Hospital (KEMH) and nearby private practices between 1989 and 1991 (https://rainestudy.org.au/)^12^. Women who had sufficient proficiency in English language, an expectation to deliver at KEMH and an intention to remain in Western Australia to facilitate the future follow-ups of the study child were eligible for the study.

Study follow-up appointments were conducted from birth and serially every two to three years during childhood and adolescence in the Raine Study Gen2 study participants. Age at follow-ups were measured to the nearest days from birth. The primary caregivers (Gen1) completed questionnaire about the study child at all follow-ups in childhood. Ethics approval for the original pregnancy cohort [Ethics approval no. DD/JS/459] and subsequent follow-ups [Ethics approval no. 382-EP_(90-23.4), 698-EP_(94-117.5), 288/EP_EC98-23.7, and 503/EP_EC00-38.2, 2019/RA/4/20/5722] were granted by the Human Research Ethics Committee of King Edward Memorial Hospital, Princess Margaret Hospital, the University of Western Australia and the Health Department of Western Australia. Parents, guardians and young adult participants provided written informed consent before enrolment of at data collection at each follow-up. This study included a subset of the original cohort that had genetic data, were Caucasian, singleton, born at term, and without evidence of fetal anomaly (n = 1328). Previous studies that examined the Raine Study Gen 2 demonstrated that the study cohort is representative of the general population in Western Australia^13,14^. All research was performed in accordance with the approved guidelines.

### Evaluating early life predictors of childhood obesity

Five early life markers were evaluated as predictors of childhood obesity at 5-, 8-, and 10-years of age. These predictors included two anthropometric variables and three genetic instruments.

#### Anthropometric variables

Two anthropometric predictors were considered: i) percent optimal birthweight (POBW), and early life BMI. POBW was expressed as a ratio of observed growth to optimal growth (adjusting for gestational age at birth, fetal sex, and maternal characteristics including age, parity and height)^15^. Gestational age (GA) was determined by either date of last menstrual period (LMP) or fetal biometry at the 18-week gestation ultrasound (USS) examination. If the difference between methods was greater than seven days, GA was derived from USS; otherwise, the LMP method was used. Birthweight was retrieved from hospital records. In this study, early life was defined as the time from birth to three years of age. Early life anthropometric measures were measured by trained research staff with participants dressed in lightweight clothing. Height was measured (to the closest 0.1cm) with participants standing in the anatomical position, palms facing forward, without shoes, heels, buttocks and head against the board using the Holtain Infantometer and Stadiometer, while weight was measured (to the nearest 100g) with the Wedderburn Chair Scales^16^. Body mass index (BMI) was calculated using measured height and weight using the formula 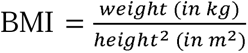. Standardised BMI Z-scores were calculated with reference to the WHO standard for age and sex^17^. In participants who attended multiple follow-ups between one- and three-years of age, data from the follow-up measured closest to three-years of age was used.

#### Genetic variables

Participants were genotyped on an Illumina 660 W Quad Array at the Centre for Applied Genomics, Toronto, Canada. Quality control (QC) of the Genome-Wide-Association-Study (GWAS) genotyped data were performed as per standard protocol. In brief, a total of 1593 participants were genotyped on an Illumina 660 Quad Array, which included 657,366 genetic variants, consisting of ∼560,000 single-nucleotide-polymorphisms (SNPs) and ∼ 95,000 copy number variants (CNVs), at the Centre for Applied Genomics, Toronto, Canada. Plate controls and replicates with a higher proportion of missing data were excluded before individuals were assessed for low genotyping success (>3% missing), excessive heterozygosity, gender discrepancies between the core data and genotyped data, and cryptic relatedness (π > 0.1875, in between second- and third-degree relatives—e.g. between half-siblings and cousins). At the SNP level, the SNP data were cleaned using PLINK^18^ following the Wellcome Trust Case—Control Consortium protocol^19^. The exclusion criteria for SNPs included: Hardy–Weinberg-Equilibrium p < 5.7 × 10–7; call-rate < 95%; minor-allele frequency < 1%; and SNPs of possible strand ambiguity (i.e. A/T and C/G SNPs). The cleaned GWAS data were imputed using MACH software^20^ across the 22-autosomes and X-chromosome against the 1000 Genome Project Phase I version 3^21^. A total of 1494 individuals with 535,632 SNPs remained after genotype QC; imputation resulted in 30,061,896 and 1,264,493 SNPs across the 22-autosomes and X-chromosome, respectively. Principal components (PCs) analysis was carried out, using SMARTPCA from v.3.0 of EIGENSOFT^22^, on the cleaned genotyped data, where PCs were generated for purposes of adjusting for population stratification in all genetic analyses.

### Development of polygenic scores (PGS)

Three polygenic scores (PGS) were examined as instruments to predict childhood obesity in this study. i) The first PGS was based on 58 single nucleotide polymorphisms (SNPs) derived from a genome wide association study of birthweight published in 2016 (BW-PGS ^23^) that was previously used as a predictor of adult obesity^24^. ii) The second PGS used 146 SNPs from the updated GWAS analyses of birthweight that was published in 2019^25^ (BW-PGS_2019_).. iii) The third PGS was developed based on 58 SNPs identified in a literature search of candidate gene and GWAS studies of childhood obesity published between 2006 and 2019 (ChOB-PGS)^26–32^.

### Childhood outcomes

This study used the childhood data collected at 5, 8, and 10-year follow-ups. Height and weight were measured as per the early life follow-up protocols described for the anthropometric predictors. Similarly, standardised BMI Z-scores were calculated based on measured height and weight and with reference to the WHO standard for age and sex^33^. Overweight was defined as being above the 85^th^ percentile or a Z-score greater than 1.0365, while obesity was defined as being above the 95^th^ percentile or a Z-score greater than 1.645.

### Statistical methods

Childhood outcomes were analysed as continuous variables unless otherwise stated. All early life predictors (anthropometric and genetic instruments) were standardised and evaluated against childhood outcomes. Univariate models, where predictors were interrogated for associations between potential predictors and childhood BMI at ages 5-, 8- and 10-years of age.

Multivariate models considered included models that accounted for both anthropometric measures and genetic instruments to examine the associations between predictors and childhood obesity. Three multivariate models were considered at each cohort review (i.e. ages 5, 8 and 10): The first model accounted for POBW, early BMI, and BW-PGS_2016_. The second model accounted for POBW, early BMI, and BW-PGS_2019_. The third model accounted for POBW, early BMI, and ChOB-PGS. (Supplementary Table 1). Additionally, the first two PCs were included in all three models to account for population stratification. Several metrics, including the coefficient of determination (R2), adjusted R2, and root mean squared error (RMSE), were computed for the assessment of model’s goodness of fit. Last, proportion of variance that explained childhood BMI were reported for predictors considered to assess their value in the fitted multivariate models. All data were analysed using R and its associated libraries. The final model for each cohort is the same and accounted for POBW, early BMI, ChOB-PGS, and the first two PCs.

## Results

### Cohort characteristics

A total of 1328 participants were available for the current analysis. Specific sample sizes for complete data analyses are included in the result tables, where applicable. Maternal and early life characteristics are summarised by sex in Table 1, while childhood anthropometric measures of obesity are summarised by sex in Table 2. Maternal age at pregnancy and anthropometric measures were similar between the sexes. The mothers of female participants had lower incidences of diabetes and hypertension, and were less likely to be nulliparous or a non-smoker when compared to their male counterparts. Except for the 8-year follow-up where female study participants were heavier, male study participants, in general, were taller and heavier, had greater weight gain in early life, and more likely to be overweight or obese at all follow-ups examined in this study.

**Table 1.** Maternal and offspring characteristics.

|  | All Study<br>Participants<br>(N=1328) | Females<br>(N=640) | Males<br>(N=688) |
| --- | --- | --- | --- |
|  | Mean (SD) or<br>N (%) | Mean (SD) or<br>N (%) | Mean (SD) or<br>N (%) |
| **Maternal Characteristics** |  |  |  |
| Age at pregnancy (in years) | 28.80 (5.79) | 28.70 (5.84) | 28.90 (5.74) |
| Completed high school<br>N (%) | 572 (43.10) | 267 (41.78) | 305 (44.33) |
| Nulliparous, N (%) | 622 (46.84) | 294 (45.94) | 328 (47.67) |
| Height (in m) | 1.64 (0.07) | 1.64 (0.07) | 1.64 (0.06) |
| Pre-pregnancy weight (in kg) | 60.68 (12.19) | 60.70 (12.30) | 60.70 (12.10) |
| Pre-pregnancy BMI (in kg/m<sup>2</sup>) | 22.53 (4.32) | 22.50 (4.28) | 22.60 (4.35) |
| Diabetes, N (%) | 42 (3.21) | 18 (2.85) | 24 (3.55) |
| Hypertension, N (%) | 336 (25.36) | 154 (24.06) | 182 (26.57) |
| Ever smoked during pregnancy, N (%) | 463 (34.86) | 236 (36.88) | 227 (32.99) |
| **Early Life Characteristics\***<br>**(N<sub>T</sub>= 1232; N<sub>F</sub>= 600; N<sub>M</sub>=632)** |  |  |  |
| Gestational age at birth (days) | 279 (8.96) | 279 (8.81) | 279 (9.10) |
| Birth Length (cm) | 49.54 (2.07) | 49.20 (1.97) | 49.90 (2.10) |
| Birth Weight (g) | 3469.45 (450.83) | 3412.00 (442.00) | 3523.00 (453.00) |
| Percent Optimal Birth Weight (POBW) | 99.00 (11.61) | 99.10 (11.50) | 98.90 (11.70) |
| Age at early life* assessment | 2.66 (0.72) | 2.67 (0.69) | 2.64 (0.73) |
| Early Life Weight (in kg) | 14.20 (2.36) | 13.90 (2.29) | 14.48 (2.39) |
| Early Life Height (in cm) | 93.10 (7.33) | 92.60 (7.01) | 93.58 (7.59) |
| WHO Z-score for early life* BMI | 0.49 (0.92) | 0.53 (0.91) | 0.45 (0.92) |
| Overweight<sup>^</sup> in early life, N (%) | 320 (25.97) | 162 (27.00) | 158 (25.00) |
| Obese<sup>**</sup> in early life, N (%) | 128 (10.39) | 60 (10.00) | 68 (10.76) |
| Birth Weight Polygenic Score (BW-PGS<sub>2016</sub><sup>†</sup>) Range: 0-116 | 65.77 (3.78) | 65.70 (3.76) | 65.80 (3.79) |
| Birth Weight Polygenic Score (BW-PGS<sub>2019</sub><sup>††</sup>) Range:0-292 | 138.9 (5.90) | 138.8 (5.92) | 138.9 (5.88) |
| Childhood Obesity Polygenic Score (ChOB-PGS<sup>^</sup>) Range:0-116 | 44.41 (5.11) | 44.29 (5.15) | 44.51 (5.08) |

N<sub>T</sub>; N<sub>F</sub>; N<sub>M</sub> are sample sizes for total, female and male participants after loss to follow-up.
\*Early life defined as the window from birth to three years of age.
<sup>^</sup>Overweight defined as WHO sex and age adjusted body mass index (BMI) Z-score $\geq 1.0365$ (equivalent to $\geq 85$ th percentile); <sup>\*\*</sup>Obesity defined as WHO sex and age adjusted BMI Z-score $\geq 1.645$ (equivalent to $\geq 95$ th percentile)
<sup>†</sup> BW-PGS<sub>2016</sub> =Birthweight polygenic score developed based on genetic variants identified through the 2016 GWAS meta-analysis of birthweight; <sup>††</sup> BW-PGS<sub>2019</sub> = Birthweight polygenic score developed based on genetic variants identified through the 2019 GWAS meta-analysis of birthweight; <sup>^</sup>ChOB-PGS = Childhood obesity polygenic score developed based on genetic variants identified through the literature search of candidate gene and GWAS of childhood obesity published between 2006 and 2019.

**Table 2.** Childhood characteristics for study participants.

|  | All Study<br>Participants<br>(N=1328) | Females<br>(N=640) | Males<br>(N=688) |
| --- | --- | --- | --- |
|  | Mean (SD) or<br>N (%) | Mean (SD) or<br>N (%) | Mean (SD) or<br>N (%) |
| **Characteristics in childhood** |  |  |  |
| **5 years of age**<br>(N<sub>T</sub>= 1165; N<sub>F</sub>= 559; N<sub>M</sub>=606) |  |  |  |
| Height at 5 years of age | 116.3 (4.68) | 115.7 (4.46) | 116.8 (4.82) |
| Weight at 5 years of age | 21.59 (3.36) | 21.34 (3.28) | 21.81 (3.42) |
| WHO Z-score for BMI at 5 years of age | 0.32 (1.03) | 0.29 (0.97) | 0.35 (1.08) |
| Overweight<sup>^</sup> at 5 years of age; N (%) | 231 (19.82) | 108 (19.32) | 123 (20.30) |
| Ever overweight<sup>^</sup> by 5 years of age, N (%) | 399 (33.28) | 178 (30.96) | 221 (35.42) |
| Obese** at 5 years of age; N (%) | 103 (8.84) | 46 (8.23) | 57 (9.41) |
| Ever having obesity** by 5 years of age, N (%) | 178 (15.10) | 76 (13.50) | 102 (16.56) |
| **8 years of age**<br>(N<sub>T</sub>= 1169; N<sub>F</sub>= 572; N<sub>M</sub>=597) |  |  |  |
| Height at 8 years of age | 129.4 (5.75) | 128.8 (5.71) | 130.1 (5.73) |
| Weight at 8 years of age | 28.53 (5.66) | 28.43 (5.68) | 28.62 (5.63) |
| WHO Z-score for BMI at 8 years of age | 0.48 (1.14) | 0.51 (1.06) | 0.45 (1.21) |
| Overweight<sup>^</sup> at 8 years of age; N (%) | 305 (26.09) | 163 (28.50) | 142 (23.79) |
| Ever overweight<sup>^</sup> by 8 years of age, N (%) | 497 (41.00) | 239 (40.43) | 258 (41.55) |
| Obese** at 8 years of age; N (%) | 172 (14.71) | 83 (14.51) | 89 (14.91) |
| Ever having obesity** by 8 years of age, N (%) | 255 (21.52) | 116 (20.07) | 139 (22.90) |
| **10 years of age**<br>(N<sub>T</sub>= 1125; N<sub>F</sub>= 535; N<sub>M</sub>=590) |  |  |  |
| Height at 10 years of age | 144.1 (6.44) | 144.2 (6.49) | 144.0 (6.40) |
| Weight at 10 years of age | 39.15 (9.00) | 39.48 (8.97) | 38.85 (9.03) |
| WHO Z-score for BMI at 10 years of age | 0.59 (1.17) | 0.56 (1.13) | 0.62 (1.21) |
| Overweight<sup>^</sup> at 10 years of age; N (%) | 372 (33.07) | 173 (32.34) | 199 (33.73) |
| Ever overweight<sup>^</sup> by 10 years of age, N (%) | 593 (49.71) | 277 (48.68) | 316 (50.64) |
| Obese** at 10 years of age; N (%) | 222 (19.73) | 105 (19.63) | 117 (19.83) |
| Ever having obesity** by 10 years of age, N (%) | 331 (28.51) | 156 (28.21) | 175 (28.78) |

N<sub>T</sub>; N<sub>F</sub>; N<sub>M</sub> are sample sizes for total, female and male participants after loss to follow-up.
<sup>^</sup>Overweight defined as WHO sex and age adjusted body mass index (BMI) Z-score $\geq 1.0365$ (equivalent to $\geq 85$ th percentile); \*\*Obese defined as WHO sex and age adjusted BMI Z-score $\geq 1.645$ (equivalent to $\geq 95$ th percentile)

### Early anthropometry predicts child BMI, and predictions are strongest in early childhood

Univariate (linear) analyses revealed POBW and early BMI positively associated with childhood BMI at ages 5, 8 and 10. (all p-values < 0.001, Table 3). In multivariable analyses, effect sizes for relationships between POBW and early BMI with child BMI were highest at age 5, and decreased through to age 10 (Table 4, Figure 1, Supplementary Tables 2-4). Additionally, early BMI explained more variance in child BMI at each age (5 years: 38.7%; 8 years: 28.4%; 10 years: 18.1%), by 5-10-fold, than other anthropometric and genetic predictors (Table 4, Supplementary Tables 2-4), after adjusting for POBW and ChOB-PGS. When comparing only anthropometric predictors, POBW explained a smaller proportion of variance in childhood BMI (age 5: 3.4%; age 8: 2.1%; age 10: 1.4%, Table 4) than early BMI, after accounting for early BMI, and ChOB-PGS.

**Figure 1.**
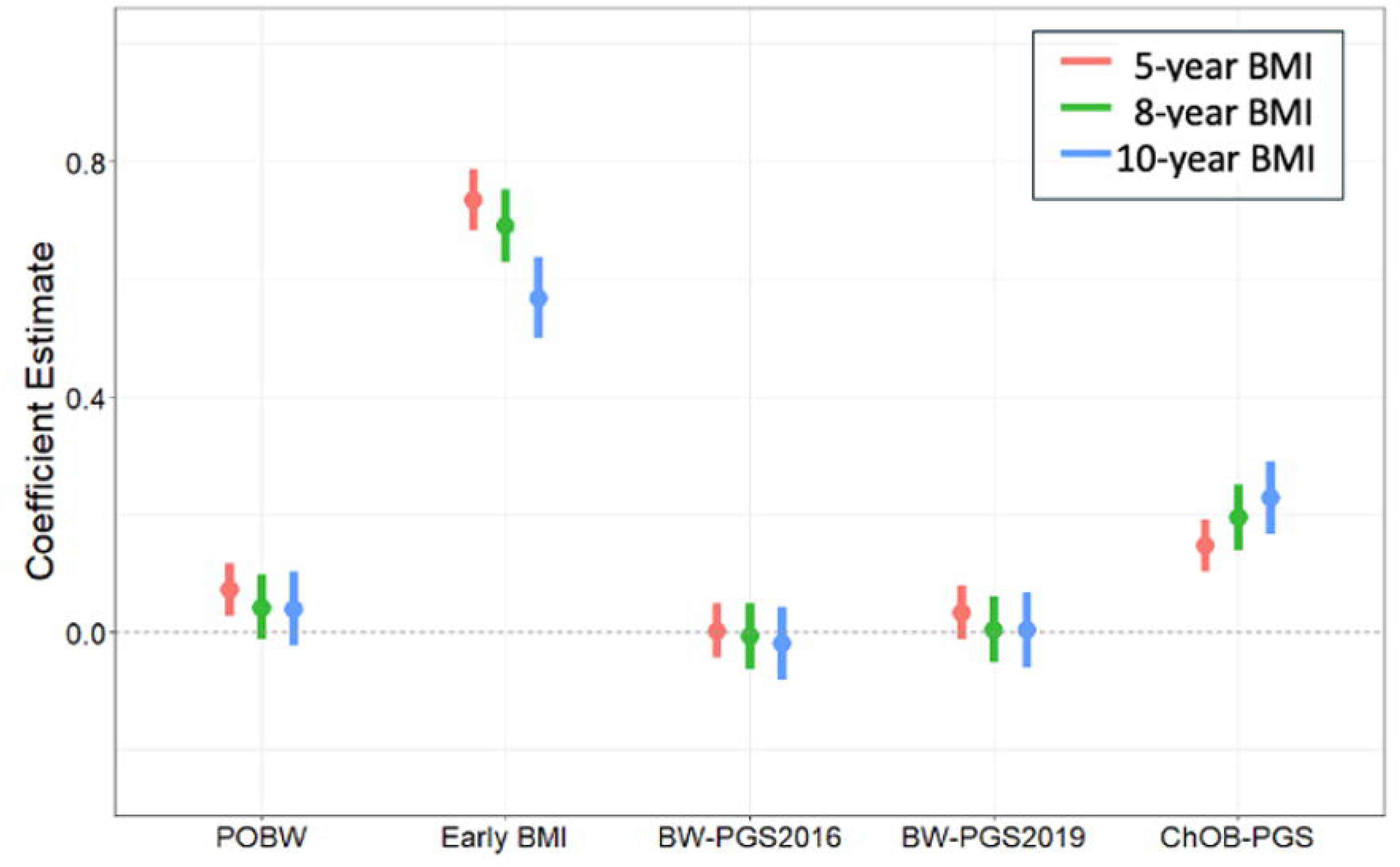
Model estimates for Childhood body mass index (BMI) models from 5- to 10-years of age. The coefficient estimate for per standard deviation increment in the predictor shown along the y-axis. The coefficient estimates for POBW, early BMI and ChOB-PGS are based on the models presented in Table 4. The coefficient estimates for BW-PGS_2016_ and BW-PGS_2019_ are based on Models 1 and 2 (described in Supplementary Table 1), respectively. ^‡^ POBW = Percent Optimal Birthweight (standardised); ^¥^ Early BMI = WHO z-standardised BMI for age and sex at in early life (based on BMI measured between birth and 3 years of age); ^†^ BW-PGS_2016_ =Birthweight polygenic score developed based on genetic variants identified through the 2016 GWAS meta-analysis of birthweight; ^††^ BW-PGS_2019_ = Birthweight polygenic score developed based on genetic variants identified through the 2019 GWAS meta-analysis of birthweight; ^^^ ChOB-PGS = Childhood obesity polygenic score developed based on genetic variants identified through the literature search of candidate gene and GWAS of childhood obesity published between 2006 and 2019. The first two principal components [^^^^PCs], derived from GWAS data of the Raine Study participants, were included in all models to account for population stratification.

**Table 3.** Univariate associations between early life predictors and childhood BMI.

|  |  | 5-year BMI |  | 8-year BMI |  | 10-year BMI |  |
| --- | --- | --- | --- | --- | --- | --- | --- |
| Class of Predictor | Predictors | Estimate (95% CI) | P | Estimate (95% CI) | P | Estimate (95% CI) | P |
| Anthropometric | POBW <sup>‡</sup> | 0.190<br>(0.131 – 0.248) | **<0.001** | 0.174<br>(0.109 – 0.240) | **<0.001** | 0.146<br>(0.078 – 0.214) | **<0.001** |
|  | Early BMI <sup>¥</sup> | 0.764<br>(0.713 – 0.815) | **<0.001** | 0.715<br>(0.654 – 0.776) | **<0.001** | 0.596<br>(0.528 – 0.665) | **<0.001** |
| Genetics | BW-PGS<sub>2016</sub> <sup>†</sup> | 0.034<br>(-0.026 – 0.094) | 0.265 | 0.034<br>(-0.032 – 0.099) | 0.315 | 0.031<br>(-0.038 – 0.100) | 0.376 |
|  | BW-PGS<sub>2019</sub> <sup>††</sup> | 0.063<br>(0.004 – 0.123) | **0.038** | 0.047<br>(-0.020 – 0.113) | 0.167 | 0.050<br>(-0.020 – 0.119) | 0.159 |
|  | ChOB-PGS <sup>^</sup> | 0.212<br>(0.154 – 0.270) | **<0.001** | 0.257<br>(0.193 – 0.321) | **<0.001** | 0.277<br>(0.210 – 0.344) | **<0.001** |

Sample sizes for 5-, 8-, 10-year BMI and POBW univariate analyses were 994, 1000, 912, respectively.
Sample sizes for 5-, 8-, 10-year BMI and Early BMI univariate analyses were 850, 842, 758, respectively
Sample sizes for 5-, 8-, 10-year BMI and Genetic univariate analyses were 1002, 1010, 922, respectively
Coefficient estimate (per SD increment) and 95% confidence interval (CI) presented for per standard deviation (SD) increment in the predictor.
Intercept coefficients not presented.
<sup>‡</sup> POBW = Percent Optimal Birthweight; <sup>¥</sup> Early body mass index (BMI) = WHO z-standardised BMI for age and sex at in early life (based on BMI measured between birth and 3 years of age); <sup>†</sup> BW-PGS<sub>2016</sub> = Birthweight polygenic score developed based on genetic variants identified through the 2016 GWAS meta-analysis of birthweight; <sup>††</sup> BW-PGS<sub>2019</sub> = Birthweight polygenic score developed based on genetic variants identified through the 2019 GWAS meta-analysis of birthweight; <sup>^</sup> ChOB-PGS = Childhood obesity polygenic score developed based on genetic variants identified through the literature search of candidate gene and GWAS of childhood obesity published between 2006 and 2019.

**Table 4.** Model summary of associations between early life predictors and childhood BMI.

| Predictors | 5-year BMI<br>(N<sub>T</sub> = 842) |  | 8-year BMI<br>(N<sub>T</sub> = 834) |  | 10-year BMI<br>(N<sub>T</sub> = 752) |  |
| --- | --- | --- | --- | --- | --- | --- |
|  | Estimate<br>(95% CI) | Percent variance<br>explained | Estimate<br>(95% CI) | Percent variance<br>explained | Estimate<br>(95% CI) | Percent variance<br>explained |
| POBW <sup>‡</sup> | **0.072**<br>**(0.026 – 0.118)** | 3.4 | 0.042<br>(-0.013 – 0.098) | 2.1 | 0.040<br>(-0.022 – 0.101) | 1.4 |
| Early BMI <sup>¥</sup> | **0.735**<br>**(0.683 – 0.786)** | 38.7 | **0.690**<br>**(0.629 – 0.752)** | 28.4 | **0.568**<br>**(0.499 – 0.637)** | 18.1 |
| ChOB-PGS <sup>^</sup> | **0.147**<br>**(0.102 – 0.192)** | 4.5 | **0.195**<br>**(0.140 – 0.249)** | 5.0 | **0.229**<br>**(0.167 – 0.290)** | 5.7 |
| Residual |  | 53.4 |  | 64.2 |  | 74.2 |
| Goodness of<br>fit metric |  |  |  |  |  |  |
| R2 | 0.47 |  | 0.36 |  | 0.26 |  |
| Adj R2 | 0.46 |  | 0.36 |  | 0.25 |  |
| RMSE | 0.76 |  | 0.91 |  | 1.01 |  |

N<sub>T</sub> denotes population sample size for complete data analysis
Multivariate association analyses for WHO z-standardised body mass index (BMI) for age and sex at 5-, 8-, and 10-year of age.
Coefficient estimate (per SD increment) and 95% confidence interval (CI) presented for per standard deviation (SD) increment in the predictor.
Intercept coefficients not presented.
<sup>‡</sup> POBW = Percent Optimal Birthweight; <sup>¥</sup> Early BMI = WHO z-standardised BMI for age and sex at in early life (based on BMI measured between birth and 3 years of age); <sup>^</sup> ChOB-PGS = Childhood obesity polygenic score developed based on genetic variants identified through the literature search of candidate gene and GWAS of childhood obesity published between 2006 and 2019. All models accounted for POBW, Early BMI, and ChOB-PGS. The first two principal components [PCs] were also included in all models to account for population stratification (estimates not presented)

### Early genetics is associated with child BMI

Across all cohort follow-ups, no significant association was observed between BW-PGS_2016_ and childhood BMI (all p-values > 0.05, Table 3) for univariate analyses. At 5, but not at 8 or 10 years of age, a positive linear association was observed between BW-PGS_2019_ and childhood BMI (p<0.05, Table 3). Positive linear associations were also reported between ChOB-PGS and childhood BMI at ages 5, 8 and 10 (all p-values < 0.001, Table 3). However, in contrast to multivariable analyses with anthropometric predictors, effect sizes for ChOB-PGS increased with age (Table 4, Figure 1, Supplementary Tables 2-4). Further, overall, approximately 5% of child BMI was explained by ChOB-PGS, with variance slightly increasing with age (Table 4).

When compared to ChOB-PGS, the proportion of variance explained by early BMI decreased from 8.66 folds at age 5, to 5.71 folds at age 8, and to 3.15 folds at age 10. The trends observed in the proportion of variance explained by predictors of childhood obesity are presented in Table 4 and Figure 2.

**Figure 2.**
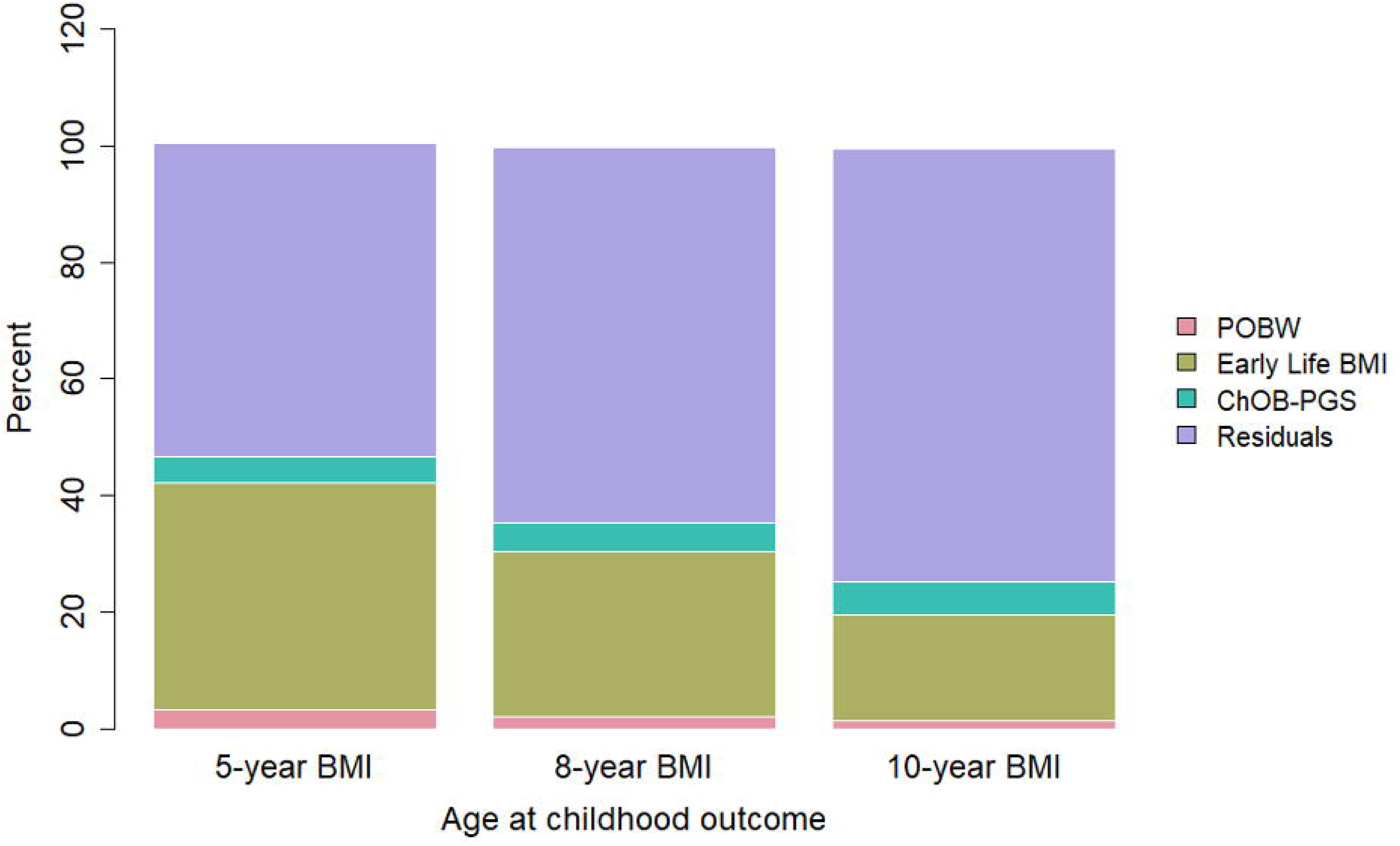
The percent of variance explained in childhood BMI by each predictor in the final multivariate models presented in Table 4. ^‡^ POBW = Percent Optimal Birthweight (standardised); ^¥^ Early BMI = WHO z-standardised BMI for age and sex at in early life (based on BMI measured between birth and 3 years of age); ^†^ BW-PGS_2016_ =Birthweight polygenic score developed based on genetic variants identified through the 2016 GWAS meta-analysis of birthweight; ^††^ BW-PGS_2019_ = Birthweight polygenic score developed based on genetic variants identified through the 2019 GWAS meta-analysis of birthweight; ^^^ ChOB-PGS = Childhood obesity polygenic score developed based on genetic variants identified through the literature search of candidate gene and GWAS of childhood obesity published between 2006 and 2019. The first two principal components [^^^^PCs], derived from GWAS data of the Raine Study participants, were included in all models to account for population stratification.

## Discussion

Here we examined two anthropometric parameters and three genetic instruments as predictors of child obesity. We identified that early BMI is a better predictor for child obesity than other parameters in later life, accounting for 10-fold more variance in BMI than the other parameters measured. These findings are significant as they point to a robust, easy measurement with clinical utility for early risk stratification of children to identify those who may most benefit from early interventions to correct BMI trajectories.

Our study demonstrates the predictive value of anthropometric measures for child obesity. Amongst the explored measures, early BMI emerged as the most robust indicator of child BMI across all ages, explaining the greatest proportion of variance at age 5, and declining by age 10. Whilst POBW was also associated with child obesity at age 5, its predictive value was lower, and not sustained over time. In predicting child obesity, early BMI, in addition to POWB, appears to enhance predictive power. Whilst growth trajectories in early life are a potential predictor of childhood obesity^34–36^, assessing these trajectories requires routine growth measurements and complex growth charts (with inconsistent agreement on which charts to use^37^, which may affect child obesity classification^38,39^). Measuring BMI at one time point in early life hence affords a simpler assessment method, increasing clinical utility. This idea is supported by recent evidence from a population study of 5173 children, that demonstrated the effect of growth trajectories did not remain significant after adjusting for BMI at 3-years, and suggested addressing childhood obesity requires BMI assessment in early life^40^. Our findings reinforce additional evidnece^34–36^ that early BMI is a potential and practical marker for risk stratification in clinical and public health settings. It should, however, be noted that the value of anthropometric measures for predicting child BMI appears to decrease with age in our cohort.

Genetic data, including BW-PGS, may strengthen models for obesity risk prediction^7^. In the current study, although genetic predictors, particularly ChOB-PGS, were associated with child BMI at all ages studied, the strength of these associations was modest compared to early BMI. ChOB-PGS explained approximately 5% of variance in BMI, with slight increase in variance explained over time, which is consistent with data that suggests the influence of genetics on BMI may increase with age, at least in adolescence^41^ and across the adult lifespan, especially in women^42^. These findings also add to the body of literature that more broadly shows age-dependent genetic effects on cardiometabolic health^43–45^. Nevertheless, our findings suggest that genetic data alone are insufficient for early life risk stratification for child obesity. Integration of anthropometric measures with genetic data (including PGS) or other environmental and lifestyle measures still holds promise for improving the precision of obesity risk prediction in childhood, as has been shown with PGSs and earlier life risk factors for myocardial infarction risk^46^. Future studies should explore combining early BMI with genetic data, particularly as the predictive strength of polygenic scores increases with age.

There are several strengths to our study. First, the Raine Study is one of the largest deeply-phenotyped and genotyped longitudinal cohorts with data collection across multiple life stages, including objective clinical assessments of weight, and high participation rate across our investigated life stages. Second, the simplicity of the parameters we examined here ensures that our findings can be explored in other replication cohorts that are likely to have similar data. One of the limitations of our study is that the Raine Study lacks follow-up data between birth and one year of age, which limits our ability to understand phenotypes at the time of adiposity peak, which occurs around 6-12 months of age, and preceding the adiposity rebound^47^. Earlier adiposity peak and rebound have been previously shown to associate with adverse adolescent^48^ and adult^49^ BMI and cardiometabolic phenotypes, and data from the Raine Study demonstrate early infant feeding can modulate adiposity rebound and BMI trajectory into adolescence^50^, pointing to early interventions for reducing later obesity risk. Future studies using data from the Raine Study and other cohorts where there is limited data during these critical windows of growth should explore ways to model longitudinal BMI trajectories accounting for these data gaps. Additionally, the relatively small sample size of the included cohorts at each age explored may limit power to detect subtle effects of genetics on child BMI. Last, whilst the Raine Study is predominantly of Caucasian ethnicity, which limits the generalisability of our findings to ethnically and racially diverse populations, our data can provide clear insights into early predictors of child obesity in similar populations.

Our findings identify early BMI as the most valuable predictor of childhood obesity, surpassing genetic and birthweight markers in its ability to account for variance in BMI. This study highlights the potential for early BMI to serve as a cornerstone for early risk stratification and targeted intervention strategies for later child obesity. While the inclusion of genetic predictors such as ChOB-PGS may provide added value, particularly as children approach adolescence, the simplicity and accessibility of early BMI make it a practical and impactful tool in combating childhood obesity.

## Data Availability

The data that support the findings of this study are available on request from the corresponding author. The data are not publicly available due to privacy and ethical reasons.

## Conflict of Interest Disclosures (includes financial disclosures)

The authors declare no conflict of interest.

## Acknowledgements

We gratefully acknowledge all Raine Study participants and their families for their continued participation in the study, as well as the Raine Study team for study co-ordination and data collection. We also thank the NHMRC and the Raine Medical Research Foundation for their support. This work was supported by resources provided by the Pawsey Supercomputing Centre with funding from the Australian Government and Government of Western Australia.

## Author Contributors

C.A.W., K.L.C. and C.E.P conceptualised and designed the study, S.J.L. and C.E.P. acquired the funds and designed the data collection and data generation, C.A.W. carried out the initial analyses, C.A.W., K.L.C. and C.E.P interpreted the results, C.A.W., K.L.C. and C.E.P drafted the initial manuscript, C.A.W., K.L.C. S.M., T.A.M. L.J.B. and C.E.P critically reviewed and revised the manuscript.

## Role of Funder/Sponsor (if any)

None of the funders had a role in the design and conduct of the study.

