## Supplementary Tables for "Early indicators of child obesity to aid future clinical trials for lifecycle obesity prevention"

Supplementary Information

Supplementary Table 1: Variables included in each model in this study

| Model | Variables |
| --- | --- |
| Model 1 | POBW, Early BMI, BW-PGS_2016_, first two PCs |
| Model 2 | POBW, Early BMI, BW-PGS_2019_, first two PCs |
| Model 3 | POBW, Early BMI, ChOB-PGS, first two PCs |

POBW = Percent Optimal Birthweight (standardised); Early BMI = WHO z-standardised BMI for age and sex at in early life (based on BMI measured between birth and 3 years of age); BW-PGS_2016_ =Birthweight polygenic score developed based on genetic variants identified through the 2016 GWAS meta-analysis of birthweight; BW-PGS_2019_ = Birthweight polygenic score developed based on genetic variants identified through the 2019 GWAS meta-analysis of birthweight; ChOB-PGS = Childhood obesity polygenic score developed based on genetic variants identified through the literature search of candidate gene and GWAS of childhood obesity published between 2006 and 2019. The first two principal components [PCs], derived from GWAS data of the Raine Study participants, were included in all models to account for population stratification.

Supplementary Table 2. Model Summary for association analyses at 5-years of age goodness of fit (N_T_ = 842)

| Predictors | Model 1 | | Model 2 | | Model 3 | |
| --- | --- | --- | --- | --- | --- | --- |
|  | Estimate  (95% CI) | Percent variance explained | Estimate  (95% CI) | Percent variance explained | Estimate  (95% CI) | Percent variance explained |
| POBW ^‡^ | **0.070**  **(0.023 – 0.117)** | 3.4 | **0.065**  **(0.018 – 0.112)** | 3.3 | **0.072**  **(0.026 – 0.118)** | 3.4 |
| Early BMI ^¥^ | **0.751**  **(0.699 – 0.803)** | 41.0 | **0.751**  **(0.699 – 0.803)** | 41.0 | **0.735**  **(0.683 – 0.786)** | 38.7 |
| BW-PGS_2016_ ^†^ | 0.010  (-0.038 – 0.057) | 0.18 |  |  |  |  |
| BW-PGS_2019_ ^††^ |  |  | 0.040  (-0.007 – 0.087) | 0.48 |  |  |
| ChOB-PGS ^^^ |  |  |  |  | **0.147**  **(0.102 – 0.192)** | 4.5 |
| Residuals |  | 55.4 |  | 55.3 |  | 53.4 |
| Goodness of fit metric |  |  |  |  |  |  |
| R2 | 0.45 | | 0.45 | | 0.47 | |
| Adj R2 | 0.44 | | 0.45 | | 0.46 | |
| RMSE | 0.77 | | 0.77 | | 0.76 | |

N_T_ denotes population sample size for complete data analysis

Multivariate association analyses for WHO z-standardised body mass index (BMI) for age and sex at 5-years of age. Coefficient estimate and 95% confidence interval (CI) presented for per standard deviation (SD) increment in the predictor. Intercept coefficients not presented. ^‡^ POBW = Percent Optimal Birthweight; ^¥^ Early BMI = WHO z-standardised BMI for age and sex at in early life (based on BMI measured between birth and 3 years of age); ^^^ ChOB-PGS = Childhood obesity polygenic score developed based on genetic variants identified through the literature search of candidate gene and GWAS of childhood obesity published between 2006 and 2019. All models are described in Supplementary Table 1. The first two principal components [PCs] were also included in all models to account for population stratification (estimates not presented)

Supplementary Table 3. Model Summary for association analyses at 8-years of age (N_T_ = 834)

| Predictors | Model 1 | | Model 2 | | Model 3 | |
| --- | --- | --- | --- | --- | --- | --- |
|  | Estimate  (95% CI) | Percent variance explained | Estimate  (95% CI) | Percent variance explained | Estimate  (95% CI) | Percent variance explained |
| POBW ^‡^ | 0.042  (-0.015 – 0.100) | 2.2 | 0.040  (-0.018 – 0.098) | 2.1 | 0.042  (-0.013 – 0.098) | 2.1 |
| Early BMI ^¥^ | **0.710**  **(0.648 – 0.773)** | 30.4 | **0.710**  **(0.647 – 0.773)** | 30.4 | **0.690**  **(0.629 – 0.752)** | 28.4 |
| BW-PGS_2016_ ^†^ | 0.003  (-0.053 – 0.059) | 0.09 |  |  |  |  |
| BW-PGS_2019_ ^††^ |  |  | 0.015  (-0.042 – 0.072) | 0.19 |  |  |
| ChOB-PGS ^^^ |  |  |  |  | **0.195**  **(0.140 – 0.249)** | 5.0 |
| Residuals |  | 67.1 |  | 67.1 |  | 64.2 |
| Goodness of fit metric |  |  |  |  |  |  |
| R2 | 0.33 | | 0.33 | | 0.36 | |
| Adj R2 | 0.33 | | 0.33 | | 0.36 | |
| RMSE | 0.93 | | 0.93 | | 0.91 | |

N_T_ denotes population sample size for complete data analysis

Multivariate association analyses for WHO z-standardised body mass index (BMI) for age and sex at 8-years of age. Coefficient estimate and 95% confidence interval (CI) presented for per standard deviation (SD) increment in the predictor. Intercept coefficients not presented. ^‡^ POBW = Percent Optimal Birthweight; ^¥^ Early BMI = WHO z-standardised BMI for age and sex at in early life (based on BMI measured between birth and 3 years of age); ^^^ ChOB-PGS = Childhood obesity polygenic score developed based on genetic variants identified through the literature search of candidate gene and GWAS of childhood obesity published between 2006 and 2019. All models are described in Supplementary Table 1. The first two principal components [PCs] were also included in all models to account for population stratification (estimates not presented)

Supplementary Table 4. Model Summary for association analyses at 10-years of age (N_T_ = 752)

| Predictors | Model 1 | | Model 2 | | Model 3 | |
| --- | --- | --- | --- | --- | --- | --- |
|  | Estimate  (95% CI) | Percent variance explained | Estimate  (95% CI) | Percent variance explained | Estimate  (95% CI) | Percent variance explained |
| POBW ^‡^ | 0.041  (-0.023 – 0.105) | 1.5 | 0.038  (-0.026 – 0.102) | 1.5 | 0.040  (-0.022 – 0.101) | 1.4 |
| Early BMI ^¥^ | **0.594**  **(0.523 – 0.664)** | 20.1 | **0.594**  **(0.523 – 0.664)** | 20.1 | **0.568**  **(0.499 – 0.637)** | 18.1 |
| BW-PGS_2016_ ^†^ | -0.004  (-0.067 – 0.060) | 0.03 |  |  |  |  |
| BW-PGS_2019_ ^††^ |  |  | 0.013  (-0.051 – 0.078) | 0.13 |  |  |
| ChOB-PGS ^^^ |  |  |  |  | **0.229**  **(0.167 – 0.290)** | 5.7 |
| Residuals |  | 78.0 |  | 78.0 |  | 74.2 |
| Goodness of fit metric |  |  |  |  |  |  |
| R2 | 0.22 | | 0.22 | | 0.26 | |
| Adj R2 | 0.22 | | 0.22 | | 0.25 | |
| RMSE | 1.0 | | 1.0 | | 1.0 | |

N_T_ denotes population sample size for complete data analysis

Multivariate association analyses for WHO z-standardised body mass index (BMI) for age and sex at 10-years of age. Coefficient estimate and 95% confidence interval (CI) presented for per standard deviation (SD) increment in the predictor. Intercept coefficients not presented. ^‡^ POBW = Percent Optimal Birthweight; ^¥^ Early BMI = WHO z-standardised BMI for age and sex at in early life (based on BMI measured between birth and 3 years of age); ^^^ ChOB-PGS = Childhood obesity polygenic score developed based on genetic variants identified through the literature search of candidate gene and GWAS of childhood obesity published between 2006 and 2019. All models are described in Supplementary Table 1. The first two principal components [PCs] were also included in all models to account for population stratification (estimates not presented)
